# Microbiota and Health Study: a prospective cohort of respiratory and diarrheal infections and associated risk factors in Bangladeshi infants under two years

**DOI:** 10.1101/19000505

**Authors:** Karine Vidal, Shamima Sultana, Alberto Prieto Patron, Aristea Binia, Mahbubur Rahman, Iztiba Mallik Deeba, Harald Brüssow, Olga Sakwinska, Shafiqul Alam Sarker

## Abstract

**Background:** Early childhood respiratory and diarrheal infections are major causes of morbidity and mortality worldwide. There is a need to further assess the epidemiology through prospective and community-based studies to gain key insights that could inform preventative measures to reduce the risk of infectious disease in this vulnerable population. We aimed to analyze the burden and determinants of acute respiratory infection (ARI) and diarrhea episodes affecting infants during their first 2 years of life with state-of-the-art molecular technologies.

**Methods:** The ARI and diarrhea episodes were prospectively collected in a community-based, longitudinal cohort of infants (n=267) from birth to 2 years of life in Bangladesh. Women were recruited during the third trimester of pregnancy. Demographic, socioeconomic, and environmental information on the households was recorded. Nasopharyngeal and fecal samples were collected during regular scheduled visits from mother-infant pairs, and also from the infants during unscheduled visits for reported illnesses. New generation sequencing methods will be utilized to determine microbiota composition and function, supplemented by the state-of-the-art multiplex molecular detection technology for a wide range of bacterial and viral pathogens.

**Discussion:** This study sought to assess the epidemiology of both respiratory and gastrointestinal illnesses during the first 2 years of life in children from a peri-urban community of Dhaka, Bangladesh. Characteristics of the mothers, as well as birth characteristics of infants enrolled in the Microbiota and Health Study are presented here. We will determine any potential association between microbiota composition and the abovementioned illnesses, and also examine the influence of known and hypothesized risk factors on the occurrence of infections. Such putative factors include environmental, socioeconomic, maternal, clinical, and selected genetic factors, namely the variation of the fucosyltransferase genes (*FUT2* and *FUT3*) of mothers and infants. This study will add to current knowledge about these early childhood infectious diseases, and will provide data to generate hypotheses for the development of nutritional approaches to be used as prevention strategies.

**Trial registration:** The study was retrospectively registered at clinicaltrials.gov as NCT02361164 (February 11, 2015).

## Background

Despite notable progress made towards improving child health in the past several decades (1, 2) infectious diseases remain highly prevalent, with respiratory and diarrheal infections representing the highest burden (3). Worldwide in 2016, diarrhea and pneumonia caused over 1.4 million deaths, and approximately 80% of these deaths occurred in children under the age of two (2). Non-specific preventive measures, such as nutrition interventions and strategies to improve water, sanitation, and hygiene have resulted in a large decline in diarrhea in children under five years of age (4). However, this decrease was less pronounced for acute respiratory infections (ARI) (5) especially in low resource settings, leading to increased importance of ARI morbidity and mortality relative to diarrhea. The worldwide estimates of ARI prevalence vary widely. According to the Demographic Health Surveys (DHS) in low and middle-income countries, the percentage of reported cases of ARI symptoms in two weeks prior to the survey in children under age of five were as high as 10.0% in Haiti in 2015-2016; 5.4% in Bangladesh in 2014 and as low as 1.4% in Armenia in 2015-16 (6). It has to be noted that the comparisons among countries are challenging, mainly due to seasonality and the fact that DHS data are not available for the same years for all the locations.

Understandably, the majority of efforts are focused on lower respiratory tract infections (7), because of their high morbidity. Yet milder upper respiratory tract infections are approximately ten-fold more frequent (8, 9), thus representing substantial burden. Approximately two thirds of children below five years of age hospitalized in Bangladesh suffered from respiratory disorders. Economic costs of ARI episodes in lower income countries is high relative to the median per capita income. For example, a multi-site study in Northern India showed that among children aged under five years, the median direct cost of ARI represented over 5% or 10% of the family annual median income depending if treated in public or private institutions respectively (10). Moreover, these estimates do not take into account the burden of increased risk of later health problems. For example, bronchiolitis in infancy was associated with increased prevalence of recurrent wheeze, asthma and allergy in early adulthood (11, 12).

Environmental risk factors for respiratory infections are well recognized, however the attributable fraction of variation is small (below 10%) [e.g. (9)], indicating that identification of other factors responsible for the differential susceptibility to respiratory infections is needed to optimize efforts aimed at prevention and treatment of ARI (2, 5). Emerging evidence indicates that commensal respiratory microbiota is an important modulator of susceptibility to respiratory infections (13, 14) as well as wheezing and asthma later in life (15). The commensal microbes could interact with respiratory pathogens in nasopharynx and modulate their infective potential (16, 17). It is also plausible that the respiratory microbiota play key roles in the immune maturation and maintenance of homeostasis as observed for gut commensals and thus impact respiratory health (16, 18). Interestingly, respiratory infections may negatively impact gut health indicating that so called gut-lung axis is bi-directional (19, 20). For example, in a model of acute influenza, airway disease affected the intestinal microbiota, resulting in higher susceptibility to bacterial pathogen invasion (21). Clinically, respiratory infections and diarrhea are often linked (22), suggesting the importance of investigating the two simultaneously (23).

To gain insights into the risk factors of ARI and diarrhea, we have conducted a prospective longitudinal study in Nandipara, a peri-urban community in Dhaka, Bangladesh. We have evaluated the role of both gastrointestinal and respiratory microbiota, including facultative pathogens, in susceptibility to ARI and diarrhea during the first two years of life. In addition, we monitored known and hypothesized risk factors, including environmental, maternal, socioeconomic, clinical and selected genetic factors.

## Methods

### Study objectives

We conducted a prospective, population-based, longitudinal birth cohort study to evaluate the association between the microbiota (nasopharyngeal and stool) and the occurrence of respiratory and gastrointestinal infections in infants during their first two years of life. To this end, we surveyed the occurrence of respiratory infections and investigated the nasopharyngeal microbiota, as well as presence of respiratory pathogens. To address the same question in relation to diarrhea, we set out to determine the association between stool microbiota, intestinal inflammation biomarkers, presence of specific intestinal pathogens, and occurrence of diarrhea. A closely linked objective was to determine the influence of other factors on the risk of infections, such as environmental, maternal and specific genetic factors, namely the variation of the fucosyltransferase genes (*FUT2* and *FUT3*) of mothers and infants.

### Study location

The study was conducted in Nandipara, a peri-urban community of Dhaka, the capital and largest city in Bangladesh (**Figure 1**). The region experiences a subtropical monsoon climate characterized by moderately warm temperatures (monthly average within 23 to 34°C during our trial) (24), high humidity (38 to 87%) and wide seasonal variations in rainfall, in three distinct seasons: a cool dry winter (November-February), a pre-monsoon hot season (March-May), and a rainy monsoon (June-October) (25). The crude birth rate in urban area is 20.8 per 1000 population (26). The community of Nandipara was chosen for its close proximity (∼8 km) to the International Center for Diarrheal Disease Research (icddr,b) campus, and presence of an iccdr,b-run health outpost with experience in supporting research activities [e.g. (27)]. Nandipara covers an area of around 4.5 km^2^ and with population density of 11500 per km^2^ (Dr Sarker, unpublished 2016 survey). All study participants lived within 4 km distance from the health outpost.

**Figure 1.**
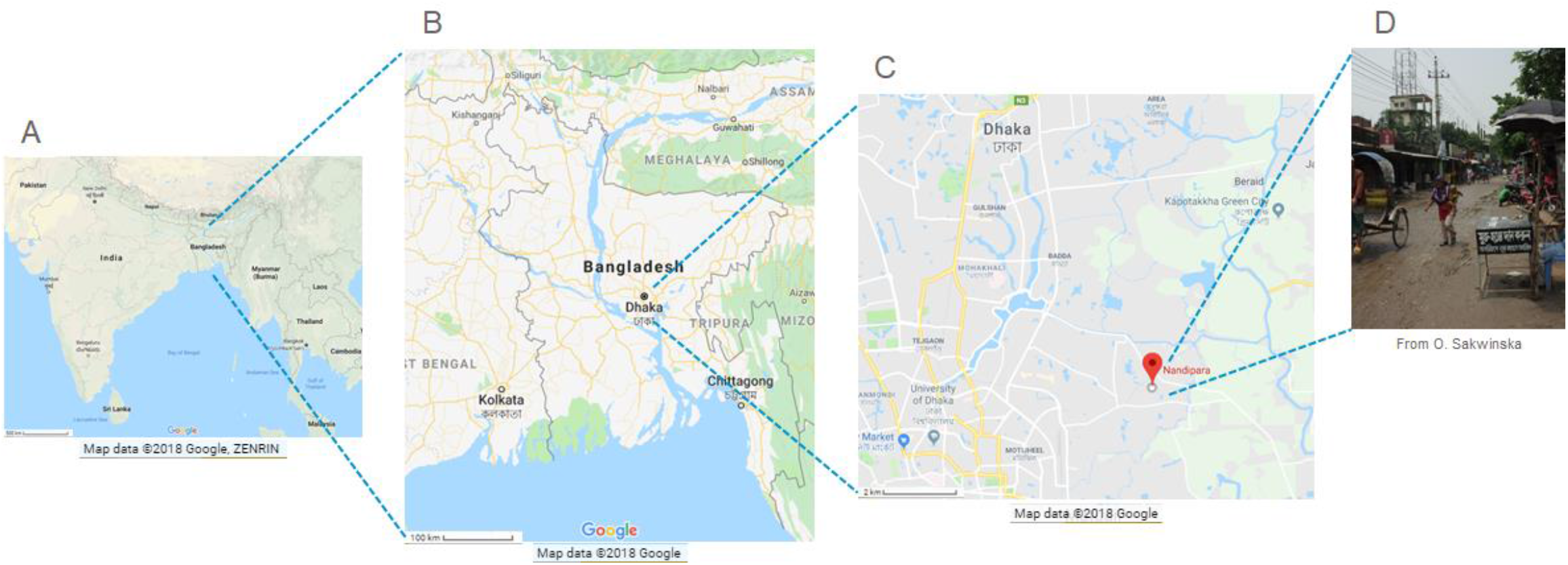
Study area. (A) Location of Bangladesh in South Asia. (B) Map of Bangladesh, with the capital Dhaka. (C) Road map showing the location of Nandipara, a peri-urban area of Dhaka. (D) Photography of Nandipara.

### Study participants

Recruitment occurred from April 2013 to October 2014; the newborn infants were followed until two years of age (*i*.*e*. until October 2016). Antenatal enrolment of women and follow-up of the study infants are shown in **Figure 2**. Pregnant women (between 16 to 28 weeks of gestation) were identified by house-to-house visits and at the icddr,b community clinic by trained field health workers and 300 women were screened. Women who fulfilled the eligibility criteria and signed the informed written consent (n=285) were enrolled at 32 ± 1 weeks of gestation. Gestational age and parity of current pregnancy was confirmed by ultrasonography. For enrolment, pregnant women needed to be inhabitants of Nandipara, between 18-35 years, with any parity but with history of healthy and uncomplicated pregnancy, and having current singleton pregnancy, healthy and uneventful progression of pregnancy and expected to have a normal pregnancy related outcome. Exclusion criteria included Rhesus negative blood group; body mass index (BMI) of less than 18.5 or more than 35, history of diabetes, hypertension; and antibiotic treatment within 3 weeks prior to this study.

**Figure 2.**
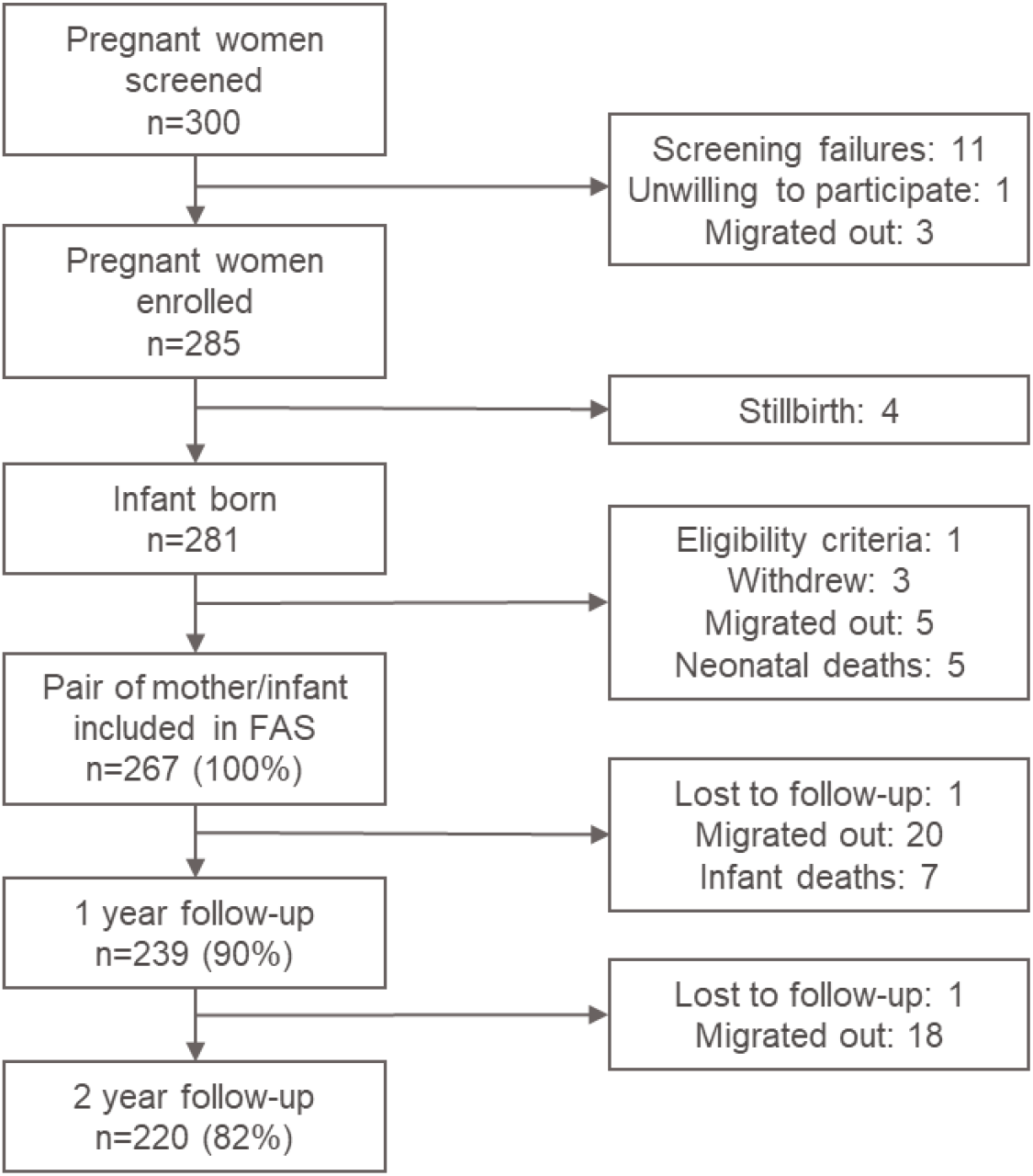
Flowchart of cohort study participants screened, enrolled and followed between April 2013 and October 2016.

### Data collection

Data and sample collection were performed by the team consisting of health workers, research assistants and study nurse supervised by a physician. Demographic, socioeconomic, environmental and mother’s health characteristics were documented via personal interview using a pre-designed epidemiological questionnaire comparable to the Bangladesh Demographic and Health Survey (BDHS). Household and mother characteristics of the study population are shown in **Table 1 and Table 2**, and birth characteristics of the infants in **Table 3**. Infant anthropometric data and feeding practice (exclusivity of breastfeeding and introduction of complementary foods) were recorded at scheduled visits, which were done at one month of age and subsequently every two months during the first year of life, and then quarterly, at 15, 18 and 24 months of age. Weight was recorded using digital weighting scale nearest 10 g (Seca, United Kingdom) and the length was measured with a manual length board to the nearest 1 cm (icddr,b-designed and validated). Mid upper arm circumference (MUAC) was recorded with a standard tape measure to the nearest 0.1 cm. In addition to the regular scheduled visits, weekly home visits were done by the study nurse supervised by a physician for the active surveillance of respiratory and gastrointestinal infections. Furthermore, mothers were asked to contact the nurse whenever her infant experienced symptoms like cough, running/blocked nose, fever, ear discharge, rapid breathing or refusal of feeding, or passage of loose stool. In such events, extra home visit was performed at the home, or mother was asked to present herself and the child to the health outpost. Each ARI symptoms were recorded as adverse events (AE) with the precise starting and ending dates. Concomitant medications (including antibiotic treatment), AE and serious adverse events (SAE) were documented in a standard electronic case report form by the medical officer.

**Table 1.** Characteristics of the households of the study participants.

| Characteristics<br>(n=267 unless stated) | Category | Frequency<br>(%) |
| --- | --- | --- |
| **Household socioeconomic characteristics** |  |  |
| **Household income (taka per month)** | <7'000<br>≥7'000 <10'000<br>>10'000 | 78 (29.2)<br>104 (39.0)<br>85 (31.8) |
| **House ownership<sup>1</sup>** | Own<br>Rent | 54 (24.5)<br>166 (75.5) |
| **Mother's education** | Illiterate<br>Primary<br>Secondary<br>High Secondary<br>University | 19 (7.1)<br>167 (62.5)<br>66 (25.7)<br>14 (5.2)<br>1 (0.4) |
| **Mother's occupation** | Housewife<br>Student<br>Working | 245 (91.8)<br>2 (0.7)<br>20 (7.5) |
| **Father's occupation** | Service<br>Business<br>Grocery shopkeeper<br>Day labor<br>Rickshaw puller<br>Auto Rickshaw driver<br>Other occupations<br>Jobless | 66 (24.7)<br>42 (15.7)<br>5 (1.9)<br>69 (25.8)<br>46 (17.2)<br>30 (11.2)<br>8 (3.0)<br>1 (0.4) |
| **Food insecurity<sup>1a</sup>** | Never<br>Rarely<br>Sometimes<br>Often | 194 (88.2)<br>22 (10.0)<br>2 (0.9)<br>2 (0.9) |
| **Household possession of goods<sup>1</sup>** | Almirah/wardrobe<br>Bed<br>Refrigerator<br>Television<br>Radio<br>Mobile phone/phone land line<br>Access to internet<sup>b</sup><br>Bicycle | 94 (42.7)<br>209 (95.0)<br>79 (35.9)<br>159 (72.3)<br>3 (1.4)<br>206 (93.6)<br>29 (13.2)<br>26 (11.8) |
| **Household size** |  |  |
| **Adults present during the day<sup>1</sup>** | 1<br>2<br>3<br>≥4 | 129 (58.6)<br>71 (32.3)<br>9 (4.1)<br>11 (5.0) |

| **Characteristics**<br>**(n=267 unless stated)** | **Category** | **Frequency**<br>**(%)** |
| --- | --- | --- |
| **Household size** |  |  |
| **Adults present at night<sup>1</sup>** | 1<br>2<br>3<br>≥4 | 4 (1.8)<br>154 (70.0)<br>18 (8.2)<br>44 (20.0) |
| **Children between 5- and 16-year old**<br>**during the day<sup>1</sup>** | 0<br>1<br>≥2 | 98 (44.5)<br>77 (35.0)<br>45 (20.5) |
| **Children between 5- and 16-year old**<br>**present at night<sup>1</sup>** | 0<br>1<br>≥2 | 96 (43.6)<br>77 (35.0)<br>47 (21.4) |
| **Children <5-year old present during**<br>**the day<sup>2</sup>** | 1<br>2<br>3 | 186 (84.9)<br>32 (14.5)<br>1 (0.5) |
| **Children <5-year old present at**<br>**night<sup>2</sup>** | 1<br>2<br>3 | 186 (84.9)<br>31 (14.2)<br>1 (0.5) |
| **Housing characteristics** |  |  |
| **Number of rooms** | 1<br>2<br>3<br>≥4 | 186 (69.7)<br>63 (23.6)<br>13 (4.9)<br>5 (1.9) |
| **Flooring material<sup>1</sup>** | Cement<br>Wood<br>Earth | 188 (85.5)<br>11 (5.0)<br>21 (9.5) |
| **Wall material<sup>1</sup>** | Tin<br>Solid<br>Mud | 123 (55.9)<br>96 (43.6)<br>1 (0.5) |
| **Roof material<sup>1</sup>** | Tin<br>Concrete<br>Plastic/mud | 178 (80.9)<br>39 (17.7)<br>3 (1.4) |
| **Household sanitation facilities** |  |  |
| **Type of toilet facility<sup>1</sup>** | Flush to piped sewer system<br>Flush to septic tank<br>Flush to pit latrine<br>Pit latrine with slab<br>Flush to somewhere else<br>Hanging toilet | 1 (0.5)<br>90 (40.9)<br>76 (34.5)<br>50 (22.7)<br>2 (0.9)<br>1 (0.5) |
| **Toilet facility shared with other**<br>**households<sup>1</sup>** | Yes<br>No | 162 (73.6)<br>58 (26.4) |
| **Hand washing** |  |  |
| **Place of hand washing<sup>1</sup>** | Inside house<br>Outside house<br>< 3 steps<br>> 3 and < 10 steps<br>> 10 steps | 35 (15.9)<br>185 (84.1)<br>26 (11.8)<br>130 (59.1)<br>29 (13.2) |
| **Hand washing after defecating<sup>1</sup>** | No<br>Yes<br>With water<br>With soap<br>With mud | 0 (0.0)<br>220 (100.0)<br>21 (9.5)<br>197 (89.5)<br>2 (0.9) |
| **Hand washing after cleaning infant**<br>**who defecated<sup>1</sup>** | No<br>Yes<br>With water<br>With soap<br>With mud | 1 (0.5)<br>219 (99.5)<br>21 (9.6)<br>196 (89.5)<br>2 (0.9) |
| **Hand washing before feeding infant<sup>1</sup>** | No<br>Yes<br>With water<br>With soap | 4 (1.8)<br>216 (98.2)<br>154 (71.3)<br>62 (28.7) |
| **Hand washing before preparing**<br>**food<sup>1</sup>** | No<br>Yes<br>With water<br>With soap | 171 (77.7)<br>49 (22.3)<br>38 (77.6)<br>11 (22.4) |
| **Hand washing before eating <sup>1</sup>** | No<br>Yes<br>With water<br>With soap | 4 (1.8)<br>216 (98.2)<br>159 (73.6)<br>57 (26.4) |
| **Household drinking water** |  |  |
| **Main source of drinking water<sup>1</sup>** | Piped into dwelling<br>Piped to yard/plot<br>Tube well<br>Dug well protected<br>Dug well unprotected<br>Surface water | 4 (1.8)<br>33 (15.0)<br>8 (3.6)<br>168 (76.4)<br>5 (2.3)<br>2 (0.9) |
| **Location of drinking water<sup>1</sup>** | Dwelling<br>Yard/plot or elsewhere | 27 (12.3)<br>193 (87.7) |
| **Water treatment prior to drinking<sup>1</sup>** | None<br>Boiled<br>Filtered | 95 (43.2)<br>117 (53.2)<br>8 (3.6) |
| **Cooking amenity** |  |  |
| **Place of cooking<sup>1</sup>** | In the house<br>In a separate building<br>Outdoors | 47 (21.4)<br>80 (36.4)<br>93 (42.3) |
| **Cooking fuel<sup>1</sup>** | Natural gas/liquefied petroleum gas<br>Wood | 209 (95.0)<br>11 (5.0) |
| **Family history of respiratory illness** |  |  |
| **Person in family with chronic cough**<br>**(>3 months/year)** | No<br>Yes | 244 (91.4)<br>23 (8.6) |
| **Smoker in family** | No<br>Yes | 89 (33.3)<br>178 (66.7) |
| <sup>1</sup> Data missing for 47 participants; <sup>2</sup> Data missing for 48 participants.<br><sup>a</sup> In the past 4 weeks, how often did you worry that your household would not have enough food?<br><sup>b</sup> In this household do you have access to internet through smart phone or computer? |  |  |

**Table 2.** Characteristics of the mothers at baseline.

| Characteristics<br>(n=267 unless stated) | Category | Frequency (%) or mean ± SD<br>(median [min, max]) |
| --- | --- | --- |
| Age (years) | | 23.7± 3.8 (23.0 [18, 35]) |
| Height (cm)<sup>1</sup> | | 149.0 ± 6.0 (148.5 [130.0, 175.5]) |
| Weight (cm) | | 52.0 ± 8.1 (50.0 [37.5, 83.0]) |
| Blood group | A+<br>AB+<br>B+<br>O+ | 66 (24.7)<br>24 (9.0)<br>90 (33.7)<br>87 (32.6) |
| Gravida | 1<br>2<br>3<br>4<br>5<br>6 | 85 (31.8)<br>88 (33.0)<br>63 (23.6)<br>23 (8.6)<br>6 (2.2)<br>2 (0.7) |
| Age of last child (years) | | 5.3 ± 2.6 (5.0 [1, 15]) |
| Place of previous delivery<sup>3</sup> | Home<br>Hospital/clinic | 125 (77.6)<br>36 (22.4) |
| Event at current labor | Normal<br>Prolonged | 246 (92.1)<br>21 (7.9) |
| <sup>1</sup> Data missing for 1 participant; <sup>2</sup> Data missing for 3 participants; <sup>3</sup> Data missing for 21 participants. |  |  |

**Table 3.** Characteristics at birth of the infants included in the birth cohort study.

| Characteristics<br>(n=267 unless stated) | Category | Frequency (%) or mean ± SD (median [min, max]) |
| --- | --- | --- |
| **Sex** | Female<br>Male | 140 (52.4)<br>127 (47.6) |
| **Gestational age** | $\geq 37$ weeks<br>$< 37$ weeks | 246 (92.1)<br>21 (7.9) |
| **Place of delivery** | Hospital/Clinic<br>Home | 120 (44.9)<br>147 (55.1) |
| **Mode of delivery** | Vaginal birth<br>Caesarean section | 199 (74.5)<br>68 (25.5) |
| **Season** | Pre-monsoon (Mar-May)<br>Rainy monsoon (Jun-Oct)<br>Cool dry winter (Nov-Feb) | 89 (33.3)<br>115 (43.1)<br>63 (23.6) |
| **Weight<sup>1</sup>** | $\geq 2,500$g<br>$< 2,500$g | 220 (83.7)<br>43 (16.3) |
| **Weight (kg)<sup>1</sup>** | | $2.8 \pm 0.4$ ($2.8$ [$1.7, 4.2$]) |
| **Length (cm)<sup>1</sup>** | | $46.9 \pm 2.0$ ($47.2$ [$41.0, 52.02$]) |
| **Number of siblings<sup>2</sup>** | 0<br>1<br>$\geq 2$ | 111 (42.0)<br>101 (38.3)<br>52 (19.7) |
| <sup>1</sup> Data missing for 4 participants; <sup>2</sup> Data missing for 3 participants. |  |  |

### Case definitions

An episode of ARI was defined as a period with one or more of the following ARI symptoms: cough, runny nose, nasal congestion, ear discharge, rapid breathing or refusal of feeding. In line with other studies (28, 29), a new episode was defined as an episode starting after 7 symptom-free days from a previous episode. Each ARI episode was further categorized as either upper or lower respiratory tract infections (URTI and LRTI, respectively). An URTI was defined as any or combination of the following respiratory symptoms in the presence or the absence of fever: cough, runny nose, nasal congestion, ear discharge, and breathing difficulty; and an LRTI was defined as the presence of cough or respiratory difficulty plus any of the following: fever, fast breathing, lower chest in-drawing, lung findings in auscultation (rales or rhonchi). A diarrhea episode was defined according to the guidelines outlined by the World Health Organization (WHO), as the passage of three or more loose or liquid stools per day. A new diarrhea episode was defined when a period of symptom-free of more than 48 hours was observed.

### Sample collection and planned analysis

At enrollment, a low vaginal swab sample was collected from each mother to assess vaginal microbiota composition. As well, a blood sample (5ml) was collected for routine biochemistry (such as serum protein and hemoglobin levels), blood group including Rhesus factor and to enable future analyses (e.g. antibody titers against specific pathogens). At delivery, a cord blood sample (5 ml) was collected for future determination of the neonate nutritional status (e.g. retinol binding protein and pre-albumin) and passive transfer of immunity from mother to infant (e.g. antibodies to *S. pneumoniae* and *H. influenzae* pathogens). Nasopharyngeal and stool samples were collected from the mother and her infant at birth or within 1-2 days post-delivery, and during scheduled visits. Whenever possible, a nasopharyngeal or stool sample was collected when infant experienced ARI or diarrhea, respectively. Nasopharyngeal samples were collected using flocked swabs for adult or pediatric swabs as per manufacturer’s instruction (Copan Diagnostics, Italy). The determination of four common facultative respiratory pathogens was performed at iccdr,b by bacterial culture using standard clinical diagnostics methods. The remaining part of the sample was frozen and stored for future analysis of viral and bacterial respiratory pathogens colonization and nasopharyngeal microbiota composition. Stool were collected for future determination of pathogen colonization, gut microbiota composition and metabolome, as well as analysis of intestinal inflammatory biomarkers. Saliva samples were collected from both the mother and the infants using Oragene DNA kits (OG-500 and OG-575, respectively) to allow for future DNA extraction and determination of *FUT2* and *FUT3* polymorphisms. All biological samples were initially stored at −20°C and subsequently transferred to −80°C for shipment on dry ice to Nestlé Research where they were stored at −80°C until analysis.

### Participant management

The enrolled mothers benefited from the antenatal medical checkups (including uro-genital health assessment, blood pressure, and blood group and hemoglobin measurements), medical advice, and basic medication such as iron and vitamin supplement, if required. Mothers with severe anemia and negative Rh factor were referred to public hospitals, including Dhaka Medical College Hospital (DMCH), Dhaka or Bangabaondhu Sheikh Mujib Medical University (BSMMU), for management and advice. Enrolled children benefited from regular follow-up of growth and development, as well as prompt, free and appropriate treatment of respiratory and diarrheal infections, such as antalgic, antihistamine, oral rehydration solution (ORS) with zinc, and antibiotics. If necessary, referral to other hospital were made. The children were vaccinated according to the UNICEF guided Expanded Program for Immunization run by the Government of Bangladesh.

### Statistical considerations

The present study is an exploratory study with a broad range of variables of interest with little or no pre-existing data available at the time the study was designed to guide a priori statistical analysis. Nevertheless, based on the clinical expertise in the target community and published information, we have attempted sample size calculation based on the incidence of ARI in early childhood and tentatively linked it to the estimates of colonization with facultative respiratory pathogens. Prevalence of colonization in the population was estimated to approximately be 30% and the average number of ARI episodes per infant was five (7, 30-32). No study disaggregated the number of ARI episodes by colonization with respiratory pathogens, we thus assumed that colonized infants suffer 25-33% more episodes than non-colonized infants. While the variation of ARI incidence was unknown, we assumed it as high as 80% of the group mean. We estimated the minimum sample size of each group to detect statistically significant difference in ARI episodes by 30% between colonized and non-colonized groups. Assuming 5% significance level and a statistical power of 90%, 225 subjects were required. With expected 10% dropout rate, 250 infants had to be included, therefore 300 pregnant mothers were recruited in this study. Data analysis was conducted using full analysis set (FAS) without imputation of missing data. The FAS includes pairs of mother and infant who satisfied criteria of target population, *i*.*e*. mothers’ eligibility criteria and infants who were born alive and have at least one post-baseline visit available. When the date of completion/discontinuation was equal to the date of last visit, the subject was classified as a dropout at this visit; if the date of completion/discontinuation was later than the date of last visit, the subject was classified as a dropout at next visit. Weight-for-age, height-for-age and weight-for-length z-scores will be calculated according to WHO child growth standards (33).

## Discussion

Here we present the study protocol of a prospective, community-based, longitudinal cohort study with detailed follow up of 267 infants during the first two years of life. This study was designed to gather data on infant health with focus on respiratory and gastrointestinal infections, collect samples from infants as well as their mothers, and assemble information about numerous potential risk factors and confounders.

### Study population

The participants of the present study inhabited peri-urban community of Nandipara. The majority of the households included in the study had monthly income higher than 7000 taka, corresponding to approximately 90 dollars or 390 international purchasing power parity (PPP) dollars between 2013 and 2016. The use of a socio-demographic questionnaire similar to the BDHS 2014 allows a direct comparison of key socioeconomic, sanitation and health indicators recorded in the present cohort with those reported at national level. It revealed that the population included in the present cohort is similar to the third and fourth wealth quintile of Bangladeshi urban households. Housing corresponds to median income families in Bangladesh, with houses built in the majority with cement floor (86%) and a single room (70%). Appropriate drinking water treatment method (57%), handwashing inside the household (16%), washing hands with soap after defecating (73%), before eating (22%) and sharing toilet facility with other households (73%) were in line with BDHS 2014 findings (26). However, some of the factors considered as important risk contributors to childhood infections, such as indoor pollution from cooking were less frequent in the current population. For example, 95% of households used gas for cooking compared to 82% using solid fuel with 50% using wood in the BDHS 2014 (26).

In this cohort study, 26% of the children were delivered by Caesarean section, which is very close to the 23% national average, and reflecting the very rapid increase from 4% in 2004 (26). According to BDHS 2014 (26), 62% of births were delivered at home, a figure similar to 55% observed in the present study.

### Study strengths

Studies of both respiratory and diarrheal infections during the first two years of life conducted in community settings are rare (23), and to our knowledge our study is the first one preformed in a low income country, particularly in combination with commensal microbiota determination. Active surveillance of infections conducted by community-based team supervised by a physician is a major strength of our study, resulting in comprehensive reporting of infections. Restricted geographic area of the surveyed community facilitated communication between families and the study team, and quality medical care received by the study participants provided further incentive to report cases of infections. In addition, the recruitment period extending over 18 months will help to disentangle the effect of the season and of infant age on the susceptibility to infections. The antenatal recruitment of pregnant women and collection of antenatal samples also constitute study strength, as it permits to assess pre-natal maternal factors known or hypothesized to contribute to the risk of childhood infections. The drop-out rate from the study was relatively low, with 90% and 82% retention at one year and two years of life, respectively. The main cause of drop-out was migration out from Nandipara, and thus unlikely to introduce major bias. Another important strength of the study is the comprehensive analysis of biological samples, including state of the art molecular detection of infectious agents, covering virus and bacteria, coupled with description of commensal microbiome, metabolome and gut health biomarkers.

### Study limitations

The recruitment was limited to one site and peri-urban settings potentially limiting the conclusions of our study, although it has to be noted that this feature allowed high quality of active surveillance of the participants. To focus on mothers and infants who were healthy at baseline and presented no specific risk factors, we excluded women younger than 18 years and with BMI below 18 recorded in the third trimester of pregnancy. However, this has likely led to exclusion of individuals with most severe health problems. Thus, when interpreting the future findings it has to be considered that the conclusions should be applied to this group and not to the general population. In the absence of chest radiography to diagnose pneumonia in this study, the distinction between upper and lower respiratory tract infections will be based on clinical assessment, potentially leading to some misclassification. Likewise, the precision of the estimate of gestational age in the present study, and resulting prematurity rate is limited. Our estimates were based on the ultrasound performed in the third trimester, as the recall of Last Menstrual Period was unreliable in this population according to the clinical practice. Nevertheless, the records of gestational age conformed to expectations (min: 32.9 weeks, max: 44.7 weeks; average: 39.5), with the exception of low prematurity rate of 8 compared to 14% WHO 2012 estimate for Bangladesh (34). However, late pregnancy enrolment precluded the observation of early preterm births, and the exclusion criteria eliminated some of the most risky pregnancies potentially explaining the observed lower rate of prematurity. In the present study, 16 of infants were classified as low birth weight (*i*.*e*. <2500 g), less than Bangladesh average of 23% reported in the National Low Birth Weight Survey conducted in 2015 (35). However, we have noted that for 14% infants, the birth weight was recorded as exactly 2500 g, suggesting inadvertent bias that commonly occurs in surveys, especially of populations with high prevalence of low birth weight (36). Future analyses will need to address this bias, as outlined by Blanc and colleagues (36).

## Data Availability

The datasets used and/or analyzed during the current study are available from the corresponding author on reasonable request.

## List of abbreviations

AE: adverse event
ARI: acute respiratory infections
BDHS: Bangladesh Demographic and Health Survey
BMI: body mass index
FAS: full analysis set
FUT: fucosyltransferase
Icddr, b: International Centre for Diarrheal Disease Research, Bangladesh
LRTI: lower respiratory tract infections
URTI: upper respiratory tract infections
WHO: World Health Organization

## Declarations

### Ethics approval and consent to participate

The local independent Institutional Review Board (IRC) comprising Research Review Committee (RRC) and Ethical Review Committee (ERC) approved the study protocol (Protocol ID: PR-12051) (September 2012), its amendments (December 2013, September 2014 and March 2017), and the related study protocol (Protocol ID: PR-15098) (December 2015). The study was designed and conducted in accordance with the declaration of Helsinki and in full conformity with the principles of the Belmont Report (April, 1979). The study was performed in compliance with the guidelines on Good Clinical Practice provided by International Conference on Harmonization/Commission on European Communities (Brussels 1991). Trained staff informed all the potential participants about the purposes, duration, procedures/methods, risks and benefits of the study, as well as the terms of confidentiality. Prior to the start of the study, all participants signed an informed written consent.

### Author’s contributions

HB, SAS and OS designed the study. SS and SAS were responsible for participant recruitment and follow up and the collection of samples. KV, PPA, AB, HB, MR, IMD and OS conducted data analysis and interpretation. KV and OS wrote the manuscript. SS, PPA, HB, and SAS contributed to writing of the manuscript. All authors read and approved the final manuscript.

## Acknowledgments

We are grateful to all the families of the infants who participated in the study, as well as the local staff, namely the field health workers, the community nurses, the research assistants (Nur Jahan Akther, Ishrat Akanda, Nasreen Sultana, and Sakina Begum) and the medical officer (Zennat Arfin) who made this study possible. We acknowledge Amelie Goyer for the clinical project management as well as Maya Shevlyakova and Pavla Kadlecova for their expert statistical support. We are also thankful to Christèle Closse for providing medical writing services. We would like to thank Nashmil Emami, Elisabeth Forbes-Blom and Armin Alaedini for helpful comments on the manuscript.

